# Fiberscopic Pattern Removal for Optimal Coverage in 3D Bladder Reconstructions of Fiberscope Cystoscopy Videos

**DOI:** 10.1101/2024.04.16.24305931

**Authors:** Rachel Eimen, Halina Krzyzanowska, Kristen R. Scarpato, Audrey K. Bowden

## Abstract

**Purpose:** In the current clinical standard of care, cystoscopic video is not routinely saved because it is cumbersome to review. Instead, clinicians rely on brief procedure notes and still frames to manage bladder pathology. Preserving discarded data via 3D reconstructions, which are convenient to review, has the potential to improve patient care. However, many clinical videos are collected by fiberscopes, which are lower cost but induce a pattern on frames that inhibits 3D reconstruction. The aim of this study is to remove the honeycomb-like pattern present in fiberscope-based cystoscopy videos to improve the quality of 3D bladder reconstructions.

**Approach:** This study introduces a novel algorithm that applies a notch filtering mask in the Fourier domain to remove the honeycomb-like pattern from clinical cystoscopy videos collected by fiberscope as a preprocessing step to 3D reconstruction. We produce 3D reconstructions with the video before and after removing the pattern, which we compare with a novel metric termed the area of reconstruction coverage (A_RC_), defined as the surface area (in pixels) of the reconstructed bladder. All statistical analyses use paired t-tests.

**Results:** Preprocessing using our method for pattern removal enabled reconstruction for all (n = 5) cystoscopy videos included in the study and produced a statistically significant increase in bladder coverage (p = 0.018).

**Conclusions:** This algorithm for pattern removal increases bladder coverage in 3D reconstructions and automates mask generation and application, which could aid implementation in time-starved clinical environments. The creation and use of 3D reconstructions can improve documentation of cystoscopic findings for future surgical navigation, thus improving patient treatment and outcomes.

## 1 Introduction

White-light cystoscopy is the gold standard for diagnosis and follow-up of bladder cancer [1–3]. Review of prior cystoscopy data is an important part of the clinical workflow for bladder cancer monitoring and treatment, in preparation for surgery, and in tracking bladder features such as tumors or sites of previous surgeries. However, full review of cystoscopy videos is cumbersome and time-consuming. Hence, the standard of care involves saving only a few frames (images) from cystoscopy videos along with accompanying textual notes, which limits the potential for comprehensive review of prior cystoscopy data.

Several groups have proposed methods to enable quick, comprehensive review of cystoscopy video data [4–7], such as 3D reconstruction, in which cystoscopy videos are converted into interactive 3D visualizations [4,5,8]. We previously developed one such 3D reconstruction pipeline termed CYSTO3D [9], which works well to produce 3D reconstructions from videos collected with the rigid cystoscopes used in operating rooms. Early tests showed, however, that CYSTO3D often fails to produce successful reconstructions from videos collected with a fiberscope: the more narrow, nimble instrument still used in many clinical cystoscopies [9]. To date, no published work shows 3D bladder reconstructions from fiberscope-collected videos. We hypothesize that the reconstruction challenges are due to the honeycomb-like fiberscope pattern that covers the view of bladder wall, inhibiting the ability of CYSTO3D and similar reconstruction pipelines to detect salient features needed to perform matching and registration across frames.

While existing methods to remove fiberscope-induced patterns exist for non-bladder applications, many invoke generalized filtering or smoothing algorithms that blur the frames [10–15] making them less useful for 3D bladder reconstruction [9] which requires more details for feature detection. Other methods involve identification of the locations of the fiber core and cladding and either require access to raw image data from the sensor [16] or only work reliably under strong, consistent imaging and lighting conditions [17–20], which cannot be guaranteed for cystoscopy videos. Less direct approaches also exist, where the primary aim is to improve image resolution and the fiberscope artifacts may be consequently removed. These methods combine multiple images taken at different positions into a single higher-resolution image [21–23] or use computational imaging and spectral coding to extract intra- and inter-fiber data that are combined into a higher-resolution image [24]. However, many of these methods do not fully remove the fiberscope artifacts, and all require additional hardware or careful manipulation of the cystoscope, both of which are difficult to implement in clinical cystoscopies.

Other promising methods selectively remove fiberscope artifacts by applying a notch filter that masks the individual peaks caused by the honeycomb-like fiberscope pattern in the frequency domain of the frame. One implementation requires *a priori* knowledge of the peak-to-peak distance in the frequency domain of the frame [25], which is often unavailable to researchers and clinicians. Two other methods also remove the artifacts by applying a notch filter [26,27], but these methods have not been applied to bladder data and do not provide guidance for selecting the filtering parameters, which can make it difficult to apply these methods to other data.

Building on these notch filter methods described above, we introduce a revised notch filtering approach to successfully remove the fiberscope pattern from cystoscopy videos while minimizing blur and loss of details. When implemented as a preprocessing step for CYSTO3D, we demonstrate that this algorithm for pattern removal generates more complete 3D reconstructions of the bladder from fiberscope cystoscopy videos than can be achieved without the preprocessing. In so doing, we present the first reconstructions generated from fiberscope cystoscopies. In what follows, we first describe the fiberscope pattern removal algorithm and then assess its utility as a preprocessing step to CYSTO3D by comparing the quality and completeness of reconstructions from fiberscope videos with and without preprocessing. We also introduce a novel metric to assess the completeness of a reconstruction, which we term the area of reconstruction coverage. This metric differs from other metrics of reconstruction completeness in that it does not require ground truth knowledge about the surface area of the scene being reconstructed [28,29].

The availability of a reliable strategy to produce 3D reconstructions of fiberscope cystoscopy videos will ultimately improve patient medical records through high quality image preservation of endoscopic findings that help inform patient care. Our approach to removing fiberscope patterns thus represents an important step for clinicians to adopt 3D reconstructions of clinical cystoscopy data to improve clinical decision-making.

## 2 Characteristics of the fiberscope pattern

To facilitate the removal of artifacts contributed by the fiberscope, we must first understand the general characteristics of the fiberscope pattern. We denote the native imaging view of a video frame acquired during standard cystoscopy as the spatial domain. In the spatial domain, one can easily observe dark circular artifacts caused by the outline of fibers present in the fiberscope (Fig. 1(A)). The fibers comprising the fiberscope are typically placed in a close-packed bundle, which causes a honeycomb-like pattern to overlay the cystoscopy frame. In the frequency domain – which is obtained by taking the fast Fourier transform of the spatial domain – the semi-regularity of the honeycomb pattern presents as a series of high-intensity peaks that are positioned at the points of a hexagon surrounding the center of the magnitude spectrum (Fig. 1(B)). The annotations in Fig. 1 guide the eye to show that the lines in the pattern correspond to peaks in the frequency domain. The orientation of the hexagon is related to the axis of alignment of the fibers in the spatial domain; this orientation can change each time the scope is reconnected to the recording tower but generally remains constant within a given cystoscopy session.

**Fig. 1.**
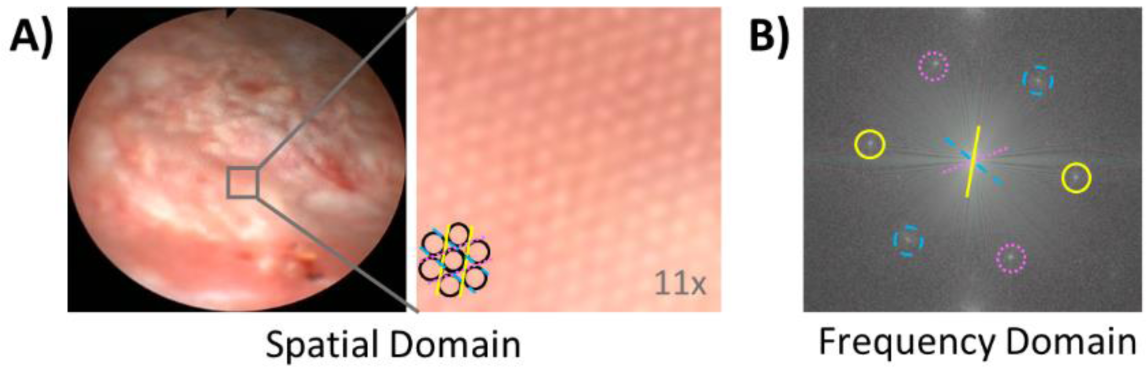
Characteristics of the pattern induced by the fiberscope, which overlays cystoscopy image content. (A) The pattern can be seen in the magnfied vew of the cystoscopy frame. Dark outlines of a few cores have been added to guide the eye in recognizing the pattern. (B) The magnitude spectrum of the frequency-domain image shows the regular orientation of the pattern, whose honeycomb packing in the spatial domain leads to distinct regions of high intensity at the points of a hexagon.

## 3 Algorithm design

Our goal was to design a pre-processing algorithm for robust 3D reconstruction of fiberscope videos with CYSTO3D. It was thus important to consider methods that would preserve contrast and minimize blur, as loss of contrast and blurring are problematic for reconstruction [29]. The algorithm comprises three general steps (Fig. 2):

1. Video cropping. We crop each frame from the video to remove the excess black border and include only the fiberscope view, which simplifies the generation of masks to remove artifacts.
2. Mask generation. We generate masks in the frequency domain to remove the fiber-induced pattern from each frame.
3. Mask application. We apply the masks as filters in the frequency domain and convert the frame back to the spatial domain.

**Fig. 2.**
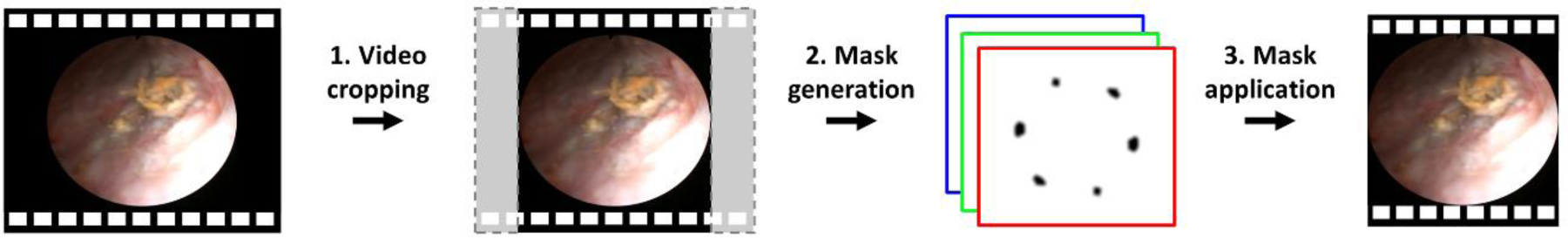
Overview of the proposed algorithm. Unprocessed cystoscopy videos are cropped, and a unique set of masks generated for individual color channels of each frame are applied in the frequency domain to produce an enhanced video with no fiberscope pattern.

### 3.1 Video cropping

The cystoscopy equipment used in this study produces frames with a native resolution of 720×480 pixels, causing the frames to present as an ellipsoidal view surrounded by black pixels. We performed a resizing operation to crop excess black pixels from the edges of the video, as many of these pixels are not equal to zero despite their irrelevance to the image and can, therefore, negatively affect mask generation. To crop the frames, we first averaged 100 randomly selected frames from the whole of the cystoscopy video to yield an “averaged frame.” We computed an averaged frame because the lighting conditions varied across frames, so the edges of the cystoscope view were not easily detectable in all frames (e.g., in dark frames where there is little contrast between the cystoscope view and the surrounding black pixels). However, the position of the cystoscope view does not change position under normal operating conditions. Thus, the averaged frame allowed us to determine the location of the cystoscope view for all frames in a video. We then identified the bounding box of the fiberscope view within this averaged frame using a boundary-detecting function in MATLAB called bwboundaries. Lastly, we cropped the individual frames to the maximum width and height of the boundary, which resulted in final frames with a resolution of approximately 480×430 pixels.

### 3.2 Mask generation

Our basic strategy to remove the fiberscope pattern from each frame involved frequency-domain filtering: that is, we applied a mask in the frequency domain of the image that removed unwanted artifacts caused by the fiberscope pattern. Notably, we expected that variations in the position of the individual red, green and blue (RGB) components of the Bayer filter would lead to slight variations in the fiberscope pattern for the different color channels; moreover, lighting varied from frame to frame and necessitated the use of a different mask for each frame. Hence, we split each frame into its individual RGB channel images prior to mask creation and converted the images to the frequency domain by performing the fast Fourier Transform. All steps of the mask-generation process were applied on a per-frame, per-channel-image basis, resulting in separate masks for each color channel of each frame.

For each color channel, we identified the fiberscope-induced peaks (FIPs) in the frequency domain using a simple thresholding operation (Sec. 3.2.1) and then performed additional processing on that initial mask, including low-pass filtering (LPF) (Sec. 3.2.2) and morphological operations (Sec. 3.2.3) to clean the peaks and minimize new artifacts caused by use of the mask. We call the final mask the “peak mask” because it removed FIPs. Fig. 3 provides an overview of the mask-generation process.

**Fig. 3.**
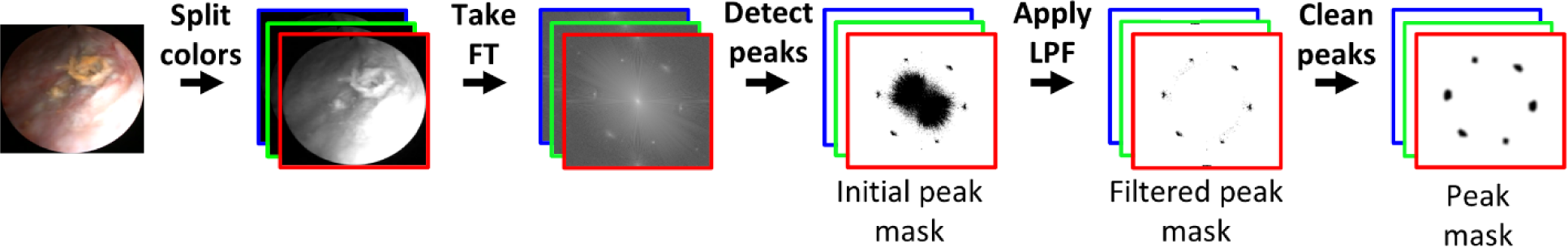
Mask generation. A cropped frame is first separated into RGB color channels. Each color channel then undergoes the following processing: fast Fourier transformation (Take FT), peak detection (Detect peaks), low-pass filtering (Apply LPF), and mask post-processing (Clean peaks), as described in Sec. 3.2.1., Sec. 3.2.2. and Sec., respectively. The end result is one peak mask for each color channel. Note, black mask content represents content that would be masked out.

#### 3.2.1 Peak detection

To detect the peaks, we performed a simple thresholding operation; the threshold used (termed the “peak detection threshold”) was a percentage of the maximum intensity of the Fourier-domain spectrum. As the mask was intended to remove unwanted content, everything above the threshold was set to zero in the mask, indicating content that would be eliminated from the final frame. When choosing a percentage to use for the threshold (Sec. 4), we found that a lower threshold degraded the frame by removing too many image features, and a higher threshold led to the detection of too few FIPs. The output mask from this process was termed our “initial peak mask.”

The use of a simple intensity threshold for peak detection, however, resulted in two undesirable effects: 1) in addition to filtering out the FIPs, it also filtered out strong image content; 2) it led to a pixelated selection of points in the area surrounding the FIPs. We addressed the former through low-pass filtering and the latter through mask post-processing.

#### 3.2.2 Low-pass filtering

While image content is found throughout the frequency spectrum, the FIPs appeared at locations of higher spatial frequency than the majority of the image content we wished to preserve. Hence, to preserve strong image content that was removed during the initial peak detection step, we modified the initial peak mask to add back in low-frequency content that had been filtered out. To do this, we created a low-pass filter (Fig. 4) from an averaged frame (generated in Sec. 2): the use of an averaged frame allowed for more reliable and consistent identification of the low-frequency components of the spectra than using data from a single frame. The filter size and shape were based on the content from the averaged frame and were determined as follows. After separating the averaged frame into three color channels (Split) and converting the color channels to the frequency domain by taking the fast Fourier transform (FT), we reconsolidated the three frames into a new grayscale frame, where the value of each pixel corresponded to the maximum of the three color-channel intensities for that pixel: that is, the channel-wise maximum (CW max.).

**Fig. 4.**
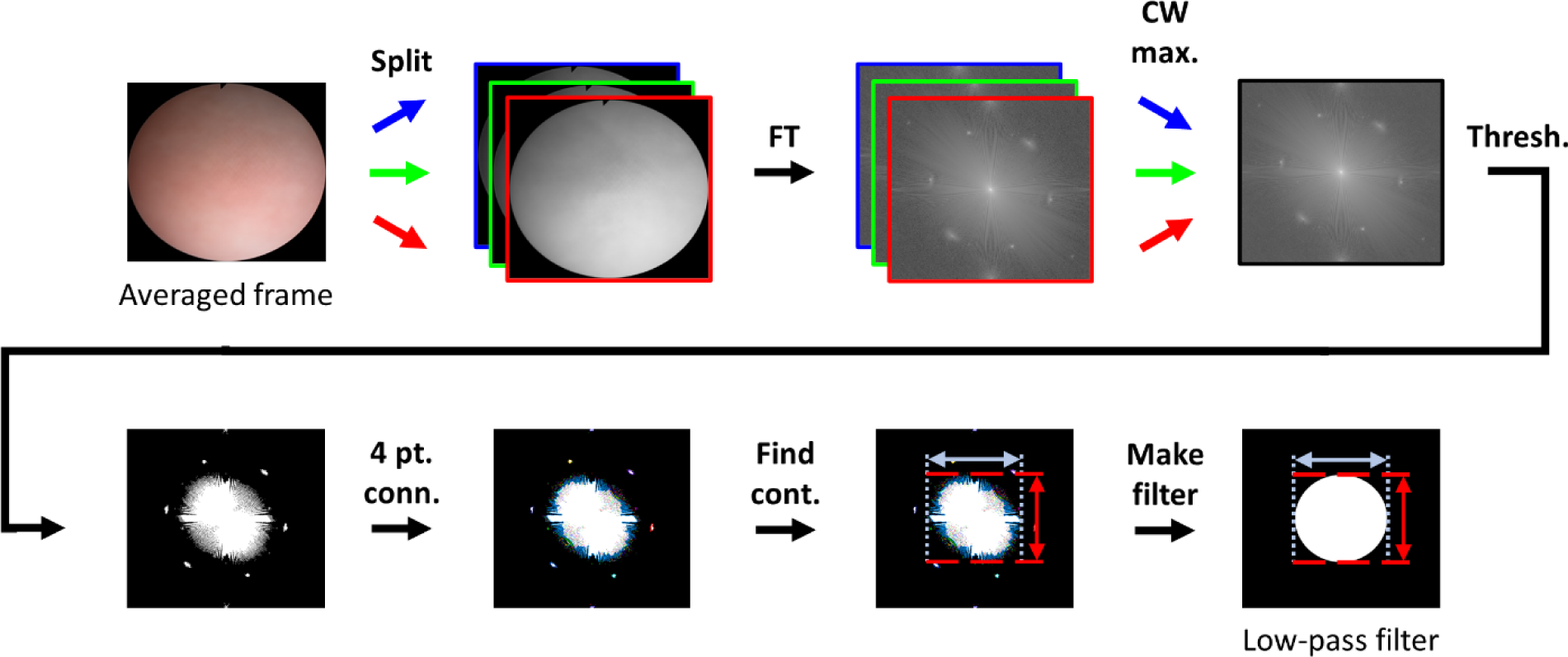
Formation of the low-pass filter. The averaged frame was separated into its color channels and converted to the frequency domain by performing a fast Fourier transform (FT). After finding the channel-wise maxima (CW max.) of the magnitude spectra, an intensity threshold was used to detect peaks in the spectra (Thresh.). We then defined contours for all regions above the thrreshold using four-point connectivity (4 pt. conn.). Lastly, the final low-pass filter was created to have the same bounds as the largest contour (Find cont.).

Next, we performed thresholding (Thresh.) at a percentage of the mean intensity of the consolidated frame, as defined in Sec. 4. In this case, content above the threshold, termed the “low-pass threshold,” was set to one, and content below the threshold was set to zero. Both FIPs and desirable image content exceeded the threshold, but we empirically determined that this threshold included sufficient low-frequency components while leaving a distinct space between the low-frequency components and the FIPs. This distinct space was important so that the low-frequency components and FIPs would not be considered part of the same contour in the next step. We then used four-point connectivity (4 pt. conn.) to detect contours (groups of connected pixels) and found the vertical and horizontal bounds of the largest contour, or the contour having the most pixels, (Find cont.). The largest contour coincided with the low-frequency image content we wished to preserve and excluded the FIPs. Lastly, we generated the final low-pass filter (Make filter) by creating an oval with the same bounds as the largest contour, where pixels inside the oval were assigned an intensity of one and pixels outside the oval an intensity of zero. The final filtering mask was ovular rather than circular because the lateral and vertical dimensions of our frame were not equal. To apply the low-pass filter to the initial peak mask generated in the previous step, we performed a logical NAND, yielding the “filtered peak mask” of Fig. 3.

#### 3.2.3 Mask post-processing

The main goal of mask post-processing (Fig. 5) was to smooth the appearance of the FIPs (to prevent new image artifacts) and to ensure that sufficient image content was preserved while removing the fiberscopic content. We first performed a morphological closing operation (kernel radius = 1) on the filtered peak mask, yielding a “closed peak mask.” This step enabled us to keep single-pixel outliers that were not attached to or associated with the FIPs but may represent image content we should preserve.

**Fig. 5.**
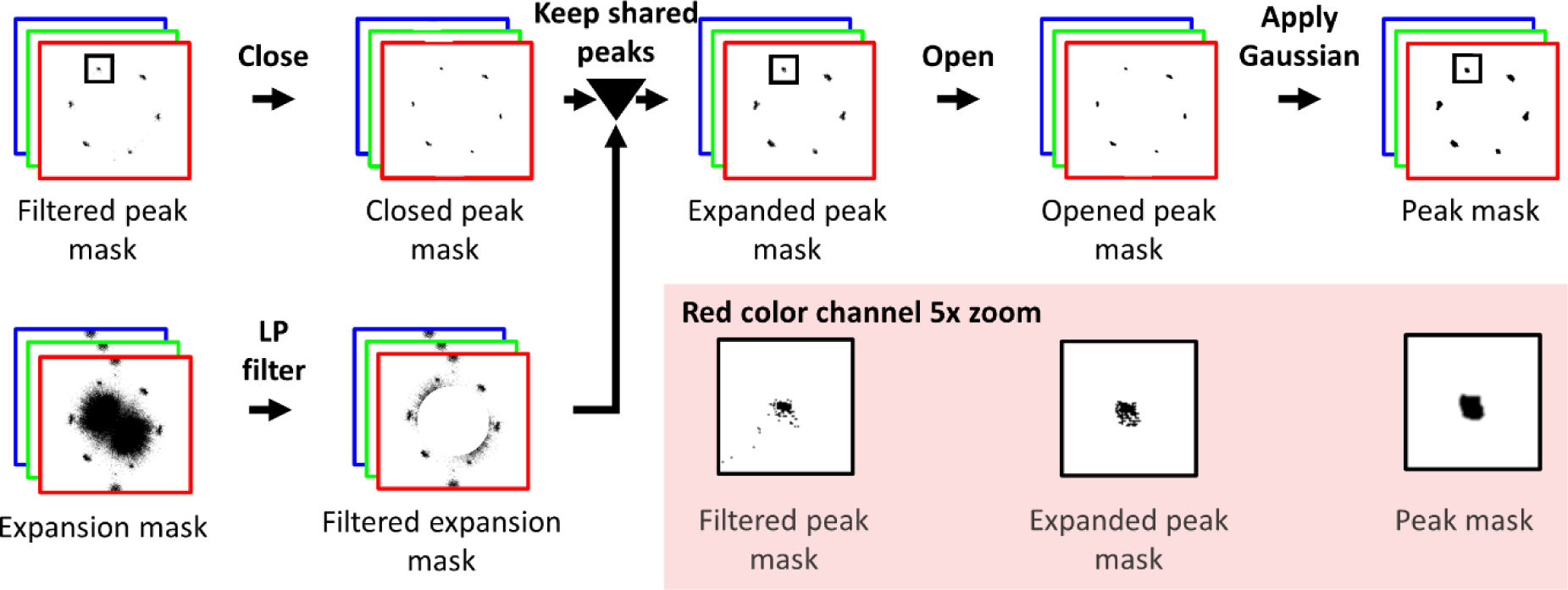
Peak mask post-processing. Morphological closing was applied to the filtered peak mask and a low-pass filter was used to remove low-frequency components from the expansion mask. Next, peaks in the filtered expansion mask that overlapped with any part of the peaks in the closed peak mask were preserved in the expanded peak mask. Morphological opening and Gaussian smoothing were then applied to produce the peak mask. Magnified regions of the masks in the red channel show the effect of select post-processing steps.

We then undertook a series of operations to enlarge the peak areas and enclose their amorphous shapes, as shown by comparing the magnified regions of the filtered peak mask and expanded peak mask. The need for the enlarged area and enclosed shapes stems from the fact that slight differences in the spacing and appearance of the fiberscope pattern throughout the frame manifested as elongated peaks with varying intensity (i.e., the FIPs are not perfect delta functions at the corners of a hexagon). In general, the threshold used to create our initial peak mask was inadequate to capture some of this elongated content, particularly because it had a lower intensity than the central peak.

Thus, we needed a way to detect these lower-intensity regions of the peaks, which resulted in enlarged peaks. To enlarge the peaks of the closed peak mask, we first created a new mask termed the “expansion mask,” which we generated by applying a threshold (termed the “peak expansion threshold”) based on the mean frame-level intensity of the spectrum. We then applied the same LPF generated in the previous step to the expansion mask, yielding the “filtered expansion mask.” We then applied the same LPF generated in the previous step to the expansion mask, yielding the “filtered expansion mask.” While the expansion mask presented larger peak regions than the closed peak mask, it also contained many outlier pixels, and the locations of the FIPs were not as clean as in the closed peak mask. We thus set out to update the closed peak mask, which contains few outlier pixels, with the enlarged areas suggested by the filtered expansion mask by examining the overlap between the closed peak mask and the expansion mask. We used a contour operation to detect groups of pixels connected with four-point connectivity in both masks. Next, we identified contours in the filtered expansion mask that overlapped with contours in the closed peak mask and updated the closed peak mask to replace its contours with those shared contours identified from the filtered expansion mask. The resulting mask was termed the “expanded peak mask.”

We then morphologically opened the expanded peak mask with a disk-shaped kernel, the diameter of which was determined in Sec. 4, with the goal of connecting the pixels within each peak of the mask. Afterwards, we applied a Gaussian smoothing filter to avoid ringing artifacts due to sharp edges on the mask. The final parameters of the smoothing filter were also chosen through the process defined in Sec. 4.

Notably, these steps resulted in a set of three final “peak masks” for each frame, one associated with each color channel from the image. As expected, there were slight differences in the appearance of the masks for each color channel. Furthermore, we verified that the masks are symmetric across their origins, which is important because they represent Fourier-domain masks for real signals, which are always symmetric across their origins.

### 3.3 Mask application

Since the masks were developed in the frequency domain, applying the mask to the frame involved simple multiplication of the color-channel-specific mask with the corresponding frequency-domain color channel of the frame. The color channels of the masked frames were recombined into a single frame, and then the inverse fast Fourier transform was used to convert back to the spatial domain, as shown in the overview of Fig 2.

## 4 Parameter selection

### 4.1 Data source

Cystoscopy videos and data from patient medical records were collected from consenting patients reporting to the Urology Clinic at Vanderbilt University Medical Center (VUMC) for a cystoscopy as part of their standard medical care. This study was approved by the Institutional Review Board (IRB) at Vanderbilt University (IRB #201269).

Cystoscopies were performed with a Karl Storz flexible cystoscope model 11272 CU1 having a field of view of 116°, a depth of field of 5-50 mm, a 22220130 NTCS camera head and an Image1 Hub 22201020 camera control unit. Overall, we collected 12 cystoscopy videos from 12 participants undergoing cystoscopies as part of their medical care. Videos were collected by two clinicians with three and 30 years of experience. Videos with large amounts of motion blur or bladder debris (n = 7) were excluded from the study because these artifacts inhibit reconstruction and are unrelated to the fiberscope pattern. We used the remaining (n = 5) videos for development and testing.

Videos ranged in duration from 30 seconds to 3 minutes (900 to 5400 frames). After each cystoscopy, we collected a video of a calibration target that comprised a grid of circles and a T-shaped alignment marker [30] from many angles and distances. Calibration videos ranged from 30 seconds to 5 minutes (900 – 9000 frames) and were used for reconstruction with CYSTO3D.

### 4.2 Selection of candidate parameter values for mask generation

We took an empirical approach to determine the parameter values (e.g., thresholds, kernel sizes) to consider for mask generation and reconstruction. First, we empirically established a range of interest for each parameter. After iterating through masks generated with the parameters in the specified ranges of interest, the parameters that resulted in proper peak coverage (that is, those that covered the FIPs without removing other data), were classified as candidate parameter values. Table 1 lists the parameters we varied, the ranges of interest we considered for each parameter and the values we classified as candidates.

**Table 1.** Parameters that were modified for mask generation along with the ranges of interest considered for each, from which we selected candidate values to use for mask generation and 3D reconstruction.

| **Parameter** | **Range of interest** | **Candidate values** |
| --- | --- | --- |
| Peak detection threshold (% max) | 45% to 65% | [ 50%, 55%, 60% ] |
| Peak expansion threshold (% mean) | 110% to 135% | [ 115%, 120%, 125, 130% ] |
| Open kernel radius | 0 to 150 | [ 10, 15 ] |
| Gaussian contour-to-kernel ratio | [ 1:6, 1:3, 1:1, 2:1 ] | [ 1:3, 1:1 ] |
| Gaussian kernel-to-sigma ratio | [1:9, 1:6, 1:3, 1:1 ] | [ 1:6, 1:3 ] |

Each threshold parameter was defined as a percentage level of the mean or maximum intensity of the frame spectrum. For the peak detection threshold, we used a percentage of the maximum intensity because the repetitive appearance of the FIPs led to strong features (high intensity) in the frequency domain. For the peak expansion threshold, we used a mean-level intensity basis because we wanted to detect the edges of the peaks, which are of lower intensity than their centers (detected with the peak detection threshold) but are of higher intensity than the mean of the spectrum. Based on early experiments, we identified that the optimal value for the low-pass threshold was 120% of the mean frame-level intensity of the averaged frame. Hence, we did not consider other candidate values for this parameter.

The candidate values for all parameters were selected by considering only those values that would lead to full peak coverage, that did not remove the low-frequency regions of the spectra, that did not connect peaks in the mask (so we could preserve data between peaks) and that smoothed the edges of the peaks. The main consideration for the open-kernel parameter was that its radius be large enough to bridge the gaps between pixels in the peaks but smaller than the peaks, as a radius larger than the peaks makes the peaks too large. The main consideration for the Gaussian kernel-to-peak ratio was to ensure that the Gaussian kernel smoothed the edges of the peaks but was equal to or smaller than the peaks, as anything larger would over-smooth the peaks. Additionally, when choosing the Gaussian sigma-to-kernel ratio, we aimed to force the Gaussian to nearly reach zero at the edges of the peaks. Most of a Gaussian area is contained within six standard deviations, so we considered a range of values around a ratio of 1:6, where higher values resulted in less FIP removal but more preservation of bladder detail, and lower values removed more of the FIPs but were more likely to induce artifacts.

The effect of parameters on mask generation is evident in Fig. 6, which shows masks for a representative spectrum (Fig. 6(A)) generated when using parameters within the list of candidate values (Fig. 6(B)) versus values in the range of interest that were not selected as candidates (Fig. 6(C)). In particular, full peak coverage was not achieved when the thresholds or Gaussian ratios were higher than the candidate values (Fig. 6(C, I, II, IV, V)) or when the open kernel diameter was set to 0 (Fig. 6(C, VIII)): that is, when no morphological opening is performed. In contrast, too many data were removed when the thresholds were lower than the candidate values (Fig. 6(C, VI – VII)) or when the open-kernel diameter was too high (Fig. 6(C, III)). Furthermore, the peaks were not smoothed, as they did not have a gradual change in intensity at their edges, when the Gaussian parameters were lower than the candidate values (Fig. 6(C, X)), which can induce artifacts in the masked frames.

**Fig. 6.**
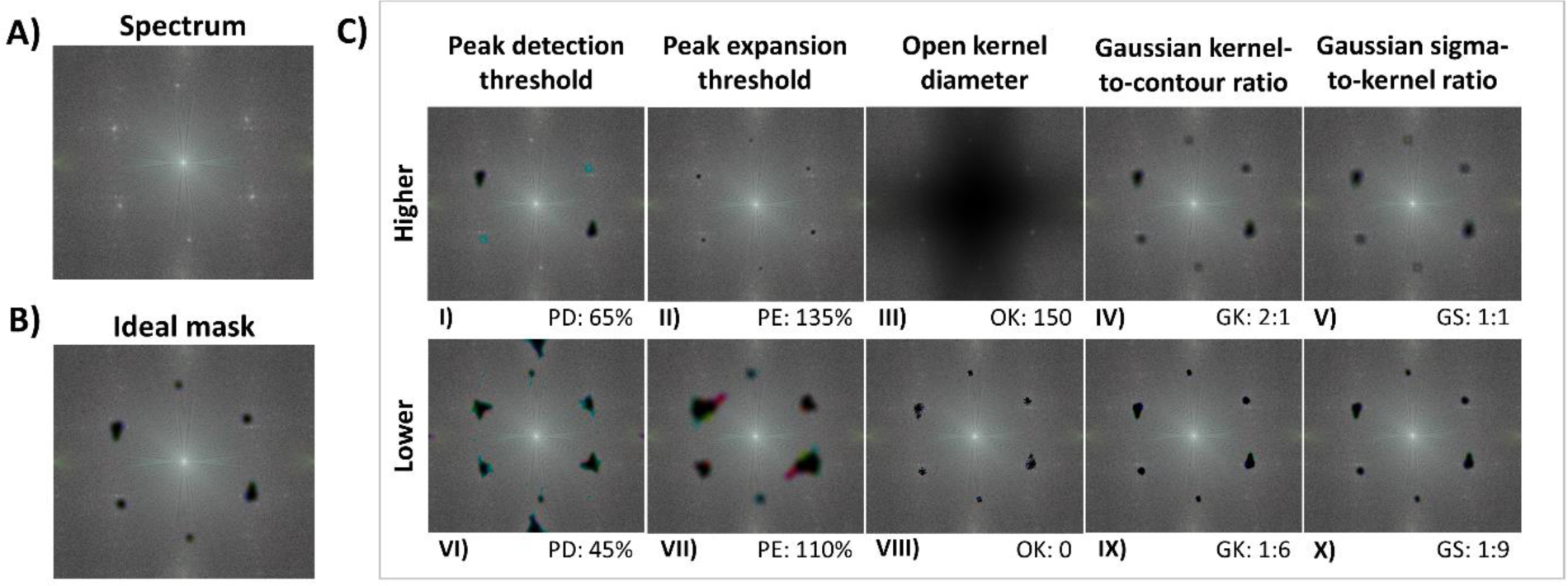
The effect of parameter values on mask generation. (A) A representative spectrum along with (B) an example of an ideal mask applied to the masked spectrum using the following parameters: peak detection threshold (PD) – 60%, peak expansion threshold (PE) – 115%, open kernel radius (OK) – 15, Gaussian kernel-to-contour ratio (GK) – 1:1, Gaussian sigma-to-kernel ratio (GS) – 1:6. The (C) masked spectra produced by changing each parameter of the ideal mask in turn. Each column contains two masked spectra where a single parameter is changed, with the top row showing the effect of an increase in the parameter and the bottom row showing the effect of a decrease. The changed parameter is listed below its respective masked spectrum.

### 4.3 Selection of optimal parameter values for mask generation

We automatically determined the optimal parameter values for mask generation from among the candidate parameters by writing a script that assessed the performance of the algorithm on 3D reconstruction outcomes and selected the parameters that resulted in the highest-quality 3D reconstruction. In brief, we compared the reconstruction performance of the unprocessed, original cystoscopy and cystoscopy videos processed with masks generated from the various candidate parameters. In total, 48 enhanced videos were generated for each cystoscopy video.

Reconstruction was performed via CYSTO3D with the parameters described previously [9], except that we used the green channel for the structure-from-motion step (as opposed to the green channel divided by the red channel), as this eliminated color channel misalignment, which was common to our fiberscopic data. It is important to note that fiberscopic pattern removal was still performed for all color channels because other steps of CYSTO3D used all three color channels. The optimal parameter values were those that yielded the most accurate and complete reconstructions relative to the unprocessed originals. Accuracy was assessed via the average reprojection error (RPE), which is a commonly used metric for 3D reconstruction assessment to measure the accuracy of the 2D-to-3D spatial mapping. Here, smaller numbers indicate better performance. Completeness was assessed with a novel metric we developed called the area of reconstruction coverage (A_RC_), which leveraged the outputs of CYSTO3D and properties of 3D reconstructions to calculate the surface area of the reconstruction in terms of pixels. There, larger numbers indicate better performance. We assessed the quality of the reconstructions in terms of their accuracy and completeness because clinicians using reconstructions prioritize both the ability to accurately determine the location of features within the bladder, such as tumors or surgical sites, and to obtain maximal coverage of the bladder surface.

In brief, the A_RC_ counts the total number of pixels from cystoscopy video frames that contribute to the final reconstruction. We chose to use pixel counts because the physical dimensions of the bladder cannot be easily extracted from the video data, and the CYSTO3D reconstruction is based on Scale Invariant Feature Transform (SIFT) [31], which also ignores scale. In contrast, the number of pixels in the reconstruction is a quantifiable measure of 3D reconstruction completeness because of the properties of the texturing pipeline [32] used by CYSTO3D. The pixel count is extracted from the texture step of the reconstruction pipeline, in which the final reconstruction is created by placing the appearance of the bladder onto the mesh (M)[32] that captures the 3D shape of the bladder. The mesh comprises a set of triangular faces (f) connecting points in 3D space; in the texture mapping step, each face of the mesh is associated with a corresponding triangular region (t_f_) of an image generated by the reconstruction pipeline (T) that contains regions of the frames in the underlying video (Fig. 7). After identifying all the image regions to be used in the reconstruction, the A_RC_ is calculated as follows:

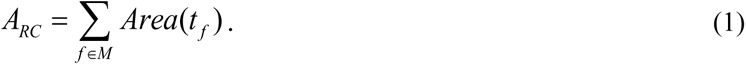

**Fig. 7.**
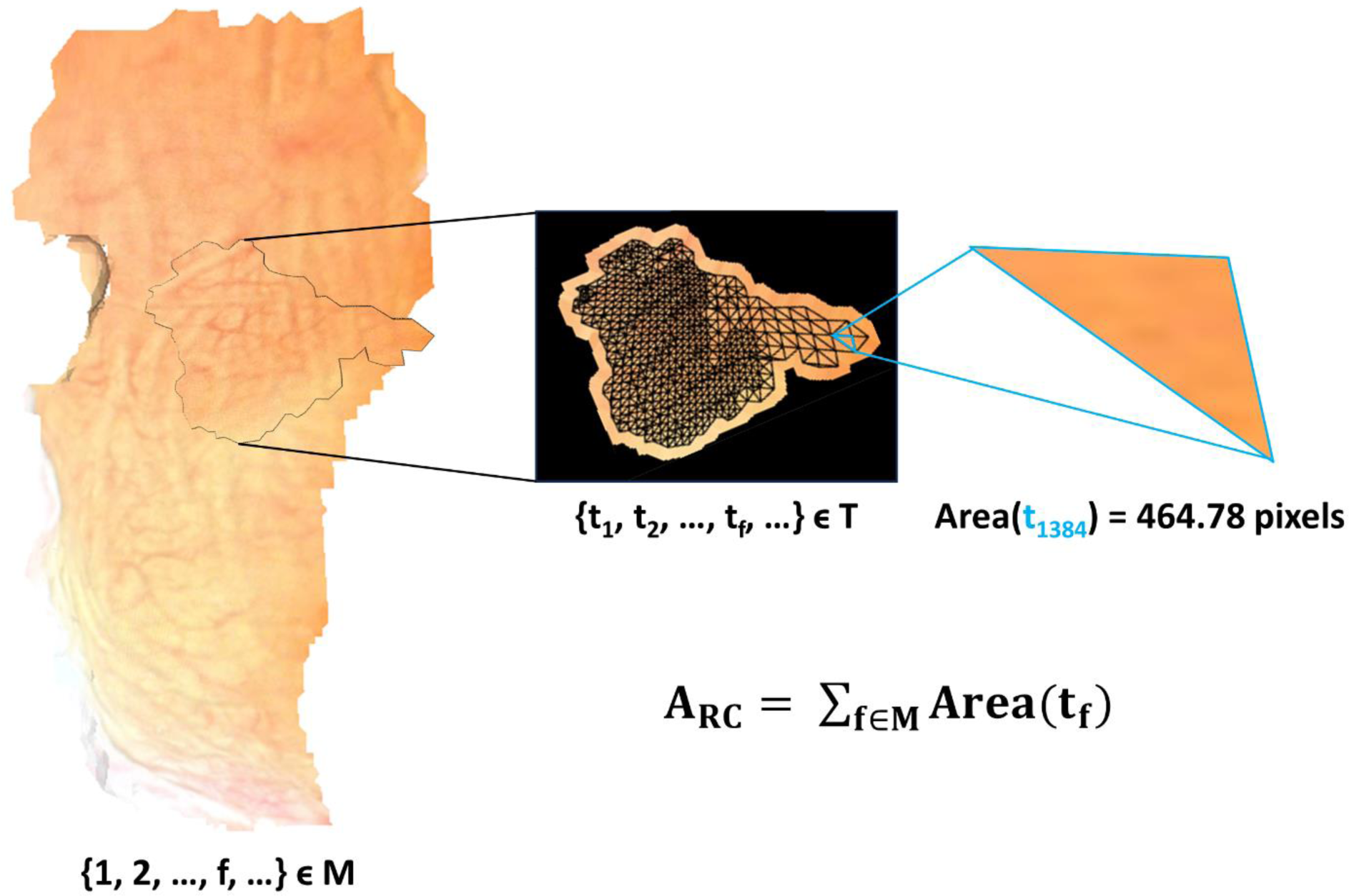
Computation of the A_RC_. The textured reconstruction comprises the mesh (M) that is covered with the texture image containing the appearance of the bladder (T), where each face (f) of M is covered with a single triangle (t_f_) in T. An example face (f = 1384) in M is covered by its corresponding triangle in T (t_f_). The area of each triangle in T is computed and added together to produce the A_RC_.

The number of pixels that cover a region of the bladder depends on the camera distance from which the frame was acquired: that is, for a given region of interest, a frame that captures the region from close-up will include more pixels than a frame that captures the region from far away. However, it is important to note that the triangular region of the frame is scaled to its corresponding triangular face on the mesh. Hence, the A_RC_ metric value is relative to the size of the mesh rather than to the distance between the camera and the bladder wall for its comprising frames. Notably, frames obtained with too far of a view are likely to be rejected by CYSTO3D as they lack sufficient features to be included in the reconstruction and frames that are close to the bladder wall are likely to be used because they will contain more distinguishable features. Thus, the A_RC_ will naturally prioritize frames that were taken at the optimal camera distance.

## 5 Results and discussion

The obtained A_RC_ and RPE values for the original video and all enhanced reconstructions generated using the candidate mask-generation parameters are shown in Fig. 8. CYSTO3D was unable to produce reconstructions for two of the five original videos, so these two reconstructions had an A_RC_ value of 0 and an undefined RPE. Notably, all reconstructions achieved an RPE less than one pixel, which indicates high accuracy; hence, our selection of optimal parameters was based exclusively on the A_RC_ values. Reconstructions for some enhanced videos gave lower A_RC_ values than their original counterparts. This is because the reconstruction performance relies on the proper balance between removing the fiberscope-induced pattern and preserving details needed for reconstruction. In cases where too many details were removed during enhancement, reconstructions with the enhanced videos can be of lower quality than their original counterparts. For all videos, there was at least one enhanced reconstruction that outperformed the original based on the A_RC_ metric. The optimal parameters for a given video ID were defined as those that produced reconstructions with the highest A_RC_ value and automatically selected.

**Fig. 8.**
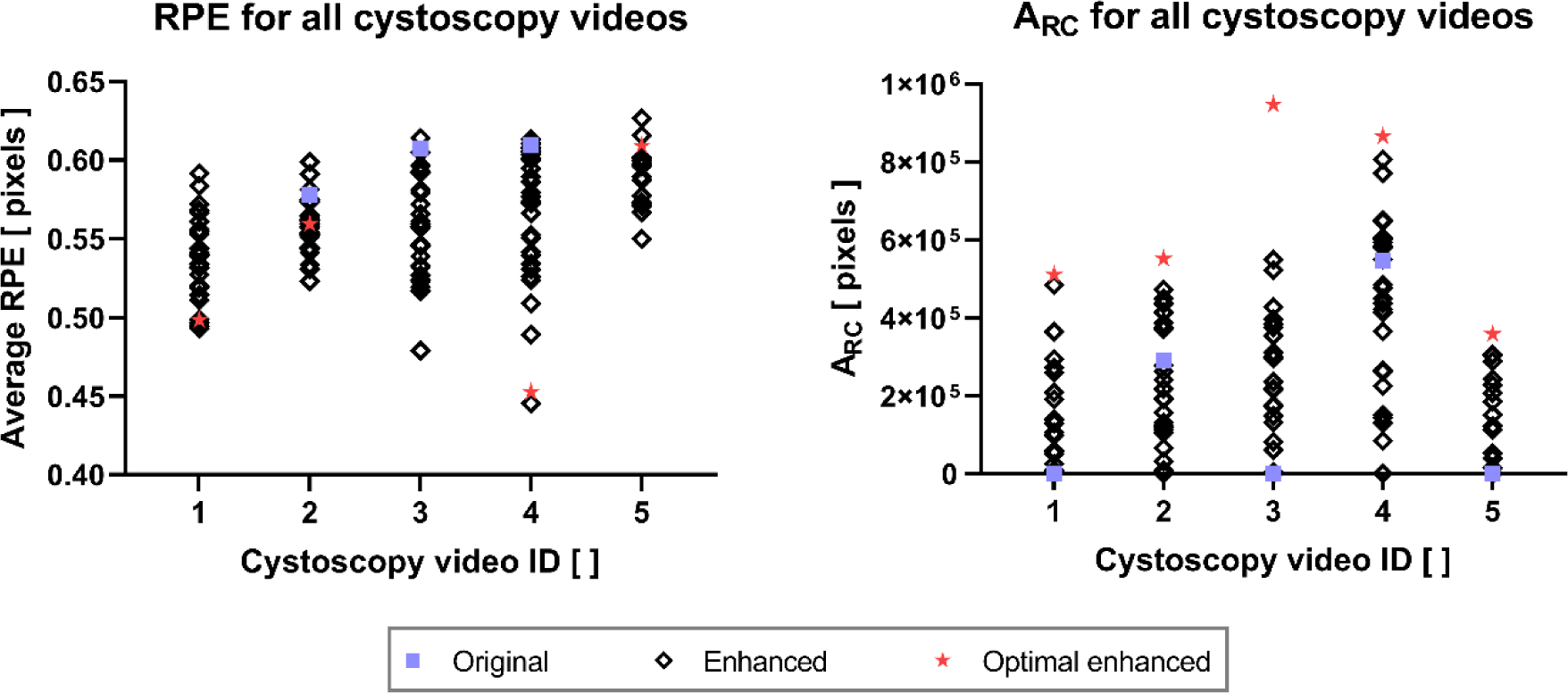
RPE and A_RC_ measurements by cystoscopy video ID. For each cystoscopy video, we compared the RPE and A_RC_ between the single original and multiple enhanced videos. Optimal enhanced videos are those that resulted in the largest A_RC_ values in their respective reconstructions.

We compared the metrics for the original and optimally enhanced reconstructions using a paired t-test (Fig. 9). Enhanced reconstructions achieved higher A_RC_ values with strong statistical significance (p = 0.018). No statistically significant change was obtained for the average RPE between the three original and enhanced video pairs, indicating that pattern removal did not affect reconstruction accuracy.

**Fig. 9.**
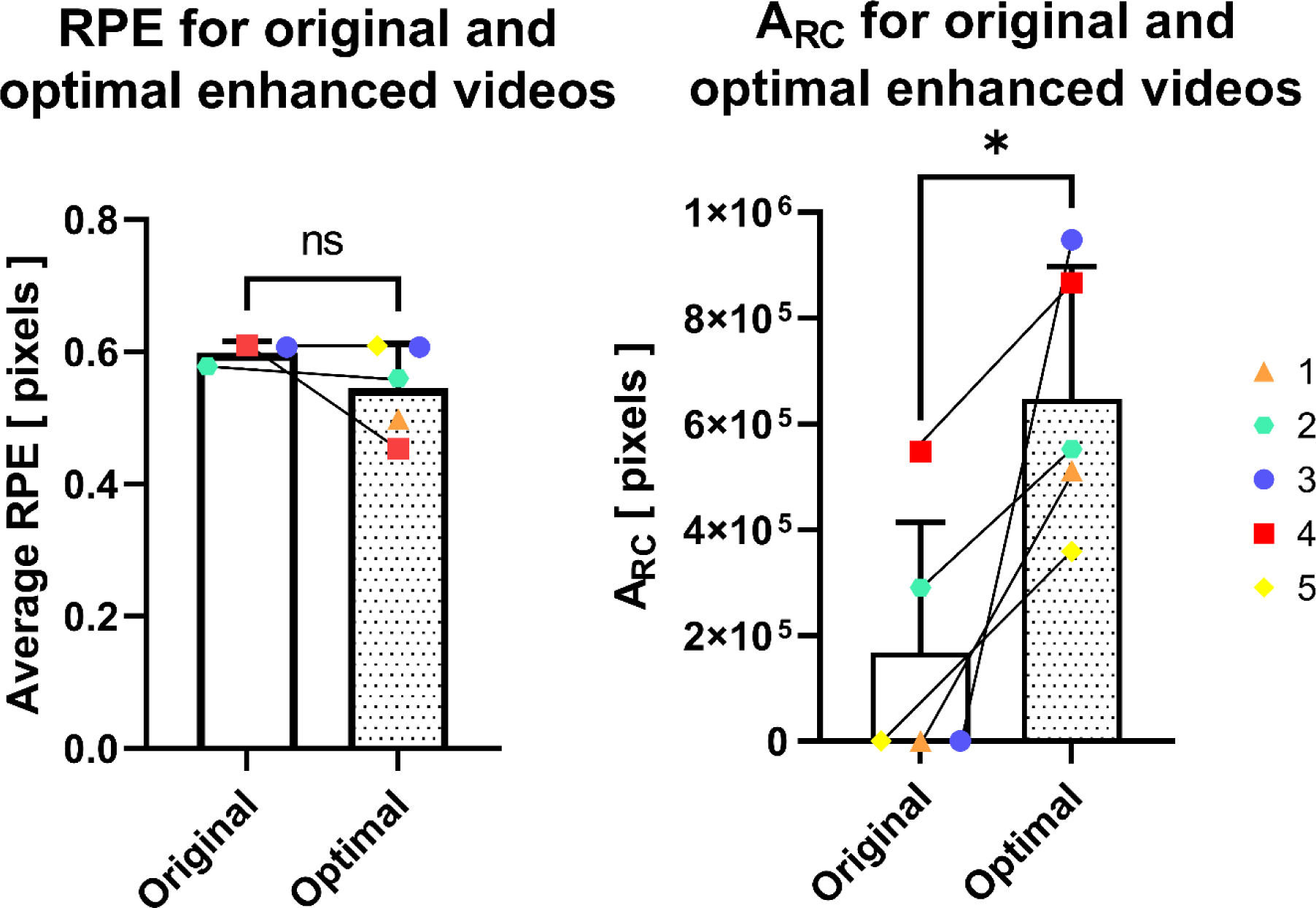
RPE and A_RC_ measurements for the original and optimal enhanced videos for each video ID. Each enhanced video used the best parameters for that particular video. In both graphs, we plotted the mean and standard deviation of the metric values for the original and optimal enhanced videos and see no statistically-significant change in the RPE (ns) and a statistically-significant increase in the A_RC_ (*).

We wrote a script that automated fiberscopic pattern removal and selected the optimal parameters for each video, where the optimal parameters were those that resulted in the largest A_RC_ metric value, as we found that the RPE values were excellent across all videos and unnecessary to improve. This script performed the following steps. First, the script performed pattern removal with every combination of parameters listed in Table 1. Second, the script initialized and ran 3D reconstructions with each of the output videos. Third, the script assessed reconstruction quality by calculating the A_RC_ metric and average RPE values for each reconstruction. Fourth, the script selected the parameters that resulted in the largest A_RC_ metric. This process was repeated for every video.

While the parameters were automatically selected, we thought it helpful to include the parameters for each video, as shown in Table 2, both to emphasize the importance of selecting unique parameters for each video and to assist in future replication of the work.

**Table 2.** Parameters that yielded the largest A_RC_ values for each video ID. The optimal parameter values that were determined include the peak detection threshold, peak expansion threshold, open kernel radius, Gaussian contour-to-kernel ratio and Gaussian kernel-to-sigma ratio.

| Video ID | Peak detection threshold | Peak expansion threshold | Open kernel radius | Gaussian contour-to-kernel ratio | Gaussian kernel-to-sigma ratio |
| --- | --- | --- | --- | --- | --- |
| 1 | 50% | 130% | 15 | 1 | 6 |
| 2 | 50% | 115% | 15 | 1 | 3 |
| 3 | 50% | 115% | 10 | 3 | 6 |
| 4 | 55% | 115% | 15 | 1 | 6 |
| 5 | 60% | 130% | 15 | 3 | 3 |

Fig. 10 further shows the reconstructions produced by CYSTO3D for both the original and optimal enhanced videos produced by the novel algorithm for pattern removal and the positive effect our algorithm has on reconstruction coverage. As the reconstructions for each video ID are shown on the same scale, it is clear that the algorithm improved the amount of recoverable content for the reconstruction. We observed that the bladder reconstructions do not recapitulate the entire bladder, and we believe this is because the collected videos contained many low-quality frames due to imaging artifacts, such as motion debris and bladder debris. These artifacts are unrelated to the fiberscopic pattern and are subjects for future improvement. Low frame quality inhibits reconstruction with CYSTO3D.

**Fig. 10.**
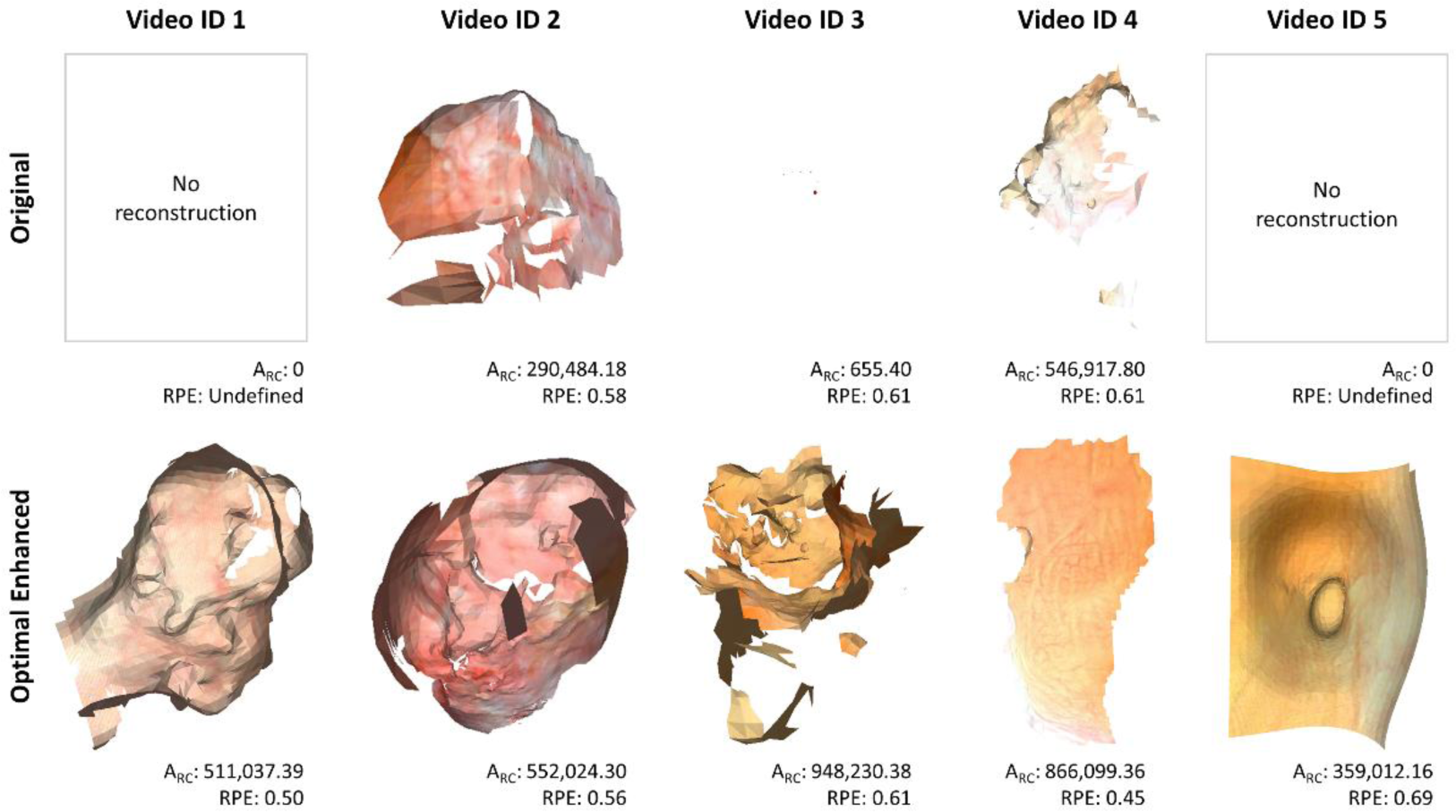
Reconstruction results with the original and optimal enhanced videos. Reconstructions with the same video ID are on the same scale, but the scales differ between video IDs.

Fig. 11 shows a visual example of a frame before and after enhancement. Notably, use of the algorithm removed prominent fiberscope features present in the original frame. Fig. 11(C) plots the intensity for each color channel from a single row in the two frames. In the original frame, high-frequency fluctuations associated with the fiberscope pattern are clearly visible, leading to a high standard deviation of the intensity of the data. In contrast, the enhanced frame data are much smoother and have a lower standard deviation (p = 0.01).

**Fig. 11.**
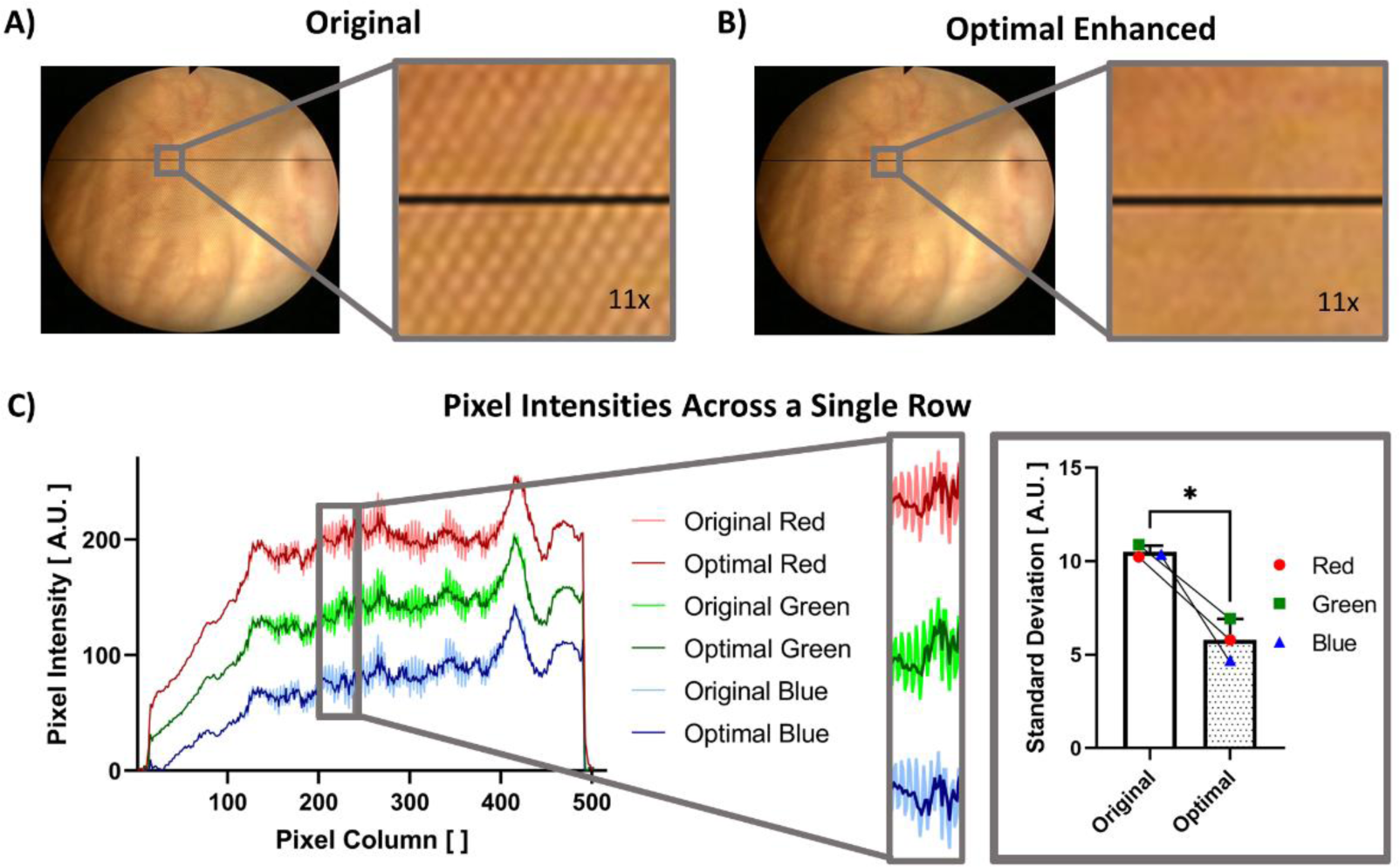
The effect of our algorithm to remove the fiberscope-induced pattern from each frame was seen through a comparison of a frame (A) before (original) and (B) after (enhanced) undergoing the algorithm, particularly in the ∼11x magnified regions of the frames. (C) The plotted pixel intensities of a single row (indicated in black on the frames) show the differences in intensity between the two frames, where the same ∼11x magnified region is shown in the plot. When observing the intensity values in this region, we also observe a statistically significant decrease in the standard deviation between the original and enhanced frames across all three color channels.

We did not develop the algorithm to remove the fiberscopic pattern or the script to calculate the A_RC_ metric in real time; rather, we focused on optimizing performance instead of decreasing runtime. The algorithm to remove the fiberscopic pattern has a runtime of 1.51 fps, while the algorithm to calculate the A_RC_ metric has a runtime of 8.59 s for a single reconstruction. Both runtimes were determined on a laptop with an Intel Core i7 processor @2.80GHz. While the runtime is not important to our application, we expect that both scripts can be optimized to have shorter runtimes in the future.

## 6 Conclusion

To our knowledge, this is the first study to produce 3D reconstructions of the bladder from fiberscopic cystoscopy videos. To achieve optimal results, we introduced a novel algorithm to detect and remove fiberscope-induced patterns while preserving details helpful to 3D reconstruction. The ability to produce 3D reconstructions from cystoscopies collected by fiberscope allows us to use cystoscopy data collected in a clinical setting to improve patient medical records and inform patient care. While digital flexible cystoscopes, which do not have fiberscope artifacts, are increasingly used in clinical settings, fiberscopes are still used around the world because of their significantly lower cost as compared to digital cystoscopes [33,34].

The methods for video cropping, peak detection and masking we use here are not specific to the appearance or size of the frame, so we anticipate they could be used for other fiberscopic equipment as well. We also expect that our algorithm could be used to remove other periodic patterns, such as patterns that appear in highly pixelated images or Moiré patterns. The Moiré pattern is a common artifact in cystoscopy videos collected by fiberscope due to interference at its detector [35,36]. We observed this pattern in some of our data and found that the FIPs have similar characteristics to the peaks in the frequency domain that are connected to the moiré pattern.

While introduction of the A_RC_ metric can objectively guide identification of the optimal reconstruction, future methods to automate the identification of candidate parameters to test or the selection of optimal parameters without testing multiple reconstructions would be beneficial. In addition, future improvements to make CYSTO3D more robust in producing larger reconstructions for a wider range of frame qualities should be pursued, as current reconstructions do not include the entire bladder because of imaging artifacts present during cystoscopic video collection.

Clinicians rely on cystoscopic data to guide clinical decision making; the ability to summarize these cystoscopy videos in a form that is informative and quick to review may optimize patient care. By removing this fiberscope pattern and producing 3D reconstructions from the enhanced videos, clinicians may quickly and easily reference cystoscopy videos collected in a variety of clinical settings, including from diagnostic office cystoscopy when fiberscopes are commonly used.

## Data Availability

All data produced in the present study are available upon reasonable request to the authors.

https://osf.io/gc9nr/

## Code, Data, and Materials Availability

The code and data described in this manuscript can be accessed through the Open Science Framework page for this paper: https://osf.io/gc9nr/

## Acknowledgements

This work was supported by members of the Vanderbilt University Medical Center Urology Clinic: Holly Marsh, Amy Luckenbaugh, Joseph Smith and Russell J. Kunic.

## References

1. H. Kobayashi, E. Kikuchi, S. Mikami, T. Maeda, N. Tanaka, A. Miyajima, K. Nakagawa, and M. Oya, "Long term follow-up in patients with initially diagnosed low grade Ta non-muscle invasive bladder tumors: Tumor recurrence and worsening progression," BMC Urol 14(1), 5 (2014).

2. K. C. Degeorge, H. R. Holt, and S. C. Hodges, Bladder Cancer: Diagnosis and Treatment (2017), 96(8).

3. M. A. Knowles and C. D. Hurst, "Molecular biology of bladder cancer: new insights into pathogenesis and clinical diversity," Nat Rev Cancer 15(1), 25–41 (2015).

4. Y. Hernández-Mier, W. C. P. M. Blondel, C. Daul, D. Wolf, and F. Guillemin, "Fast construction of panoramic images for cystoscopic exploration," Computerized Medical Imaging and Graphics 34(7), 579–592 (2010).

5. T. D. Soper, M. P. Porter, and E. J. Seibel, "Surface mosaics of the bladder reconstructed from endoscopic video for automated surveillance," IEEE Trans Biomed Eng 59(6), 1670–1680 (2012).

6. R. Miranda-Luna, C. Daul, W. C. P. M. Blondel, Y. Hernandez-Mier, D. Wolf, and F. Guillemin, "Mosaicing of bladder endoscopic image sequences: Distortion calibration and registration algorithm," IEEE Trans Biomed Eng 55(2), 541–553 (2008).

7. A. Behrens, M. Bommes, T. Stehle, S. Gross, S. Leonhardt, and T. Aach, "Real-time image composition of bladder mosaics in fluorescence endoscopy," in Computer Science - Research and Development (2011), 26(1–2), pp. 51–64.

8. R. Miranda-Luna, C. Daul, W. C. P. M. Blondel, Y. Hernandez-Mier, D. Wolf, and F. Guillemin, "Mosaicing of bladder endoscopic image sequences: Distortion calibration and registration algorithm," IEEE Trans Biomed Eng 55(2), 541–553 (2008).

9. K. L. Lurie, R. Angst, D. V. Zlatev, J. C. Liao, and A. K. Ellerbee Bowden, "3D reconstruction of cystoscopy videos for comprehensive bladder records," Biomed Opt Express 8(4), 2106 (2017).

10. D. G. Kirsch, H. L. Fu, J. Q. Brown, J. Mueller, M. J. Whitley, N. Ramanujam, R. Willett, and R. Chitalia, "Algorithms for differentiating between images of heterogeneous tissue across fluorescence microscopes," Biomedical Optics Express, Vol. 7, Issue 9, pp. 3412-3424 7(9), 3412–3424 (2016).

11. M. Dumripatanachod and W. Piyawattanametha, "A fast depixelation method of fiber bundle image for an embedded system," BMEiCON 2015 - 8th Biomedical Engineering International Conference (2016).

12. A. Shinde and M. Vadakke Matham, "Pixelate Removal in an Image Fiber Probe Endoscope Incorporating Comb Structure Removal Methods," Article in Journal of Medical Imaging and Health Informatics 4, 203–211 (2014).

13. M. Suter, J. Reinhardt, P. Montague, P. Taft, J. Lee, and J. Zabner, "Bronchoscopic imaging of pulmonary mucosal vasculature responses to inflammatory mediators," J Biomed Opt 10(3), 034013 (2005).

14. C. Winter, S. Rupp, M. Elter, C. Münzenmayer, H. Gerhäuser, and T. Wittenberg, "Automatic adaptive enhancement for images obtained with fiberscopic endoscopes," IEEE Trans Biomed Eng 53(10), 2035–2046 (2006).

15. M. M. Dickens, D. J. Bornhop, and S. Mitra, "Removal of Optical Fiber Interference in Color Micro-endoscopic Images," in Eleventh IEEE Symposium on Computer-Based Medical Systems (1998), pp. 246–251.

16. C. Winter, T. Zerfaß, M. Elter, S. Rupp, and T. Wittenberg, "Physically Motivated Enhancement of Color Images for Fiber Endoscopy," in MICCAI (2007), pp. 360–367.

17. S. Rupp, M. Elter, and C. Winter, "Improving the accuracy of feature extraction for flexible endoscope calibration by spatial super resolution.," Conference proceedings : … Annual International Conference of the IEEE Engineering in Medicine and Biology Society. IEEE Engineering in Medicine and Biology Society. Conference 6566–6572 (2007).

18. O. Icasio-Hernández, J. J. Gonzalez-Barbosa, J. B. Hurtado-Ramos, and M. Viliesid-Alonso, "3D reconstruction of hollow parts analyzing images acquired by a fiberscope," Meas Sci Technol 25(7), 075402 (2014).

19. T. N. Ford, D. Lim, and J. Mertz, "Fast optically sectioned fluorescence HiLo endomicroscopy," J Biomed Opt 17(2), 021105 (2012).

20. M. Elter, S. Rupp, and C. Winter, "Physically Motivated Reconstruction of Fiberscopic Images," in 18th International Conference on Pattern Recognition (ICPR 2006) (2006), pp. 599–602.

21. C. Renteria, J. Suárez, A. Licudine, and S. A. Boppart, "Depixelation and enhancement of fiber bundle images by bundle rotation," Appl Opt 59(2), 536 (2020).

22. Y. Huang, W. Zhou, B. Xu, J. Liu, D. Xiong, and X. Yang, "Resolution improvement in real-time and video mosaicing for fiber bundle imaging," OSA Contin 4(10), 2577 (2021).

23. J. Shao, J. Zhang, R. Liang, and K. Barnard, "Fiber bundle imaging resolution enhancement using deep learning," Opt Express 27(11), 15880 (2019).

24. J. P. Dumas, M. A. Lodhi, W. U. Bajwa, and M. C. Pierce, "Computational imaging with spectral coding increases the spatial resolution of fiber optic bundles," Opt Lett 48(5), 1088 (2023).

25. C.-Y. Lee and J.-H. Han, "Integrated spatio-spectral method for efficiently suppressing honeycomb pattern artifact in imaging fiber bundle microscopy," Opt Commun 306, 67– 73 (2013).

26. B. Regeling, B. Thies, A. O. H. Gerstner, S. Westermann, N. A. Müller, J. Bendix, and W. Laffers, "Hyperspectral Imaging Using Flexible Endoscopy for Laryngeal Cancer Detection," Sensors 16, (2016).

27. I. Aizenberg and C. Butakoff, "A windowed Gaussian notch filter for quasi-periodic noise removal," Image Vis Comput 26, 1347–1353 (2008).

28. X. Zhao, R. Wu, Z. Zhou, and W. Wu, "A new metric for measuring image-based 3D reconstruction | IEEE Conference Publication | IEEE Xplore," in 21st International Conference on Pattern Recognition (2012), pp. 1030–1033.

29. Y. Zhou, R. L. Eimen, E. J. Seibel, and A. K. Bowden, "Cost-Efficient Video Synthesis and Evaluation for Development of Virtual 3D Endoscopy," IEEE J Transl Eng Health Med 9, (2021).

30. C. Wengert, M. Reeff, P. C. Cattin, and G. Székely, "Fully Automatic Endoscope Calibration for Intraoperative Use," Bildverarbeitung für die Medizin 2006 419–423 (2006).

31. D. G. Lowe, "Distinctive image features from scale-invariant keypoints," Int J Comput Vis 60(2), 91–110 (2004).

32. M. Waechter, N. Moehrle, and M. Goesele, "Let there be color! Large-scale texturing of 3D reconstructions," Lecture Notes in Computer Science (including subseries Lecture Notes in Artificial Intelligence and Lecture Notes in Bioinformatics) 8693 **LNCS**(PART 5), 836–850 (2014).

33. U. Micoogullari and V. Ulker, "The Comparison of the Image Quality of Portable Miniature and Conventional Light Sources Used in Flexible Cystoscopy: An In Vitro Evaluation.," Journal of Urological Surgery 9(1), 47–52 (2022).

34. S. S. Quayle, C. D. Ames, D. Lieber, Y. Yan, and J. Landman, "COMPARISON OF OPTICAL RESOLUTION WITH DIGITAL AND STANDARD FIBEROPTIC CYSTOSCOPES IN AN IN VITRO MODEL," Urology 66(3), 489–493 (2005).

35. M. Monga and E. A. Klein, eds., Ureteroscopy (Springer Science+Business Media, 2013).

36. A. C. F. Ng, M. Y. C. Wong, and S. Isotani, eds., Practical Management of Urinary Stone (Springer, 2021).

